# Unveiling Women’s Experiences Amidst the COVID-19 Pandemic in Nigeria: An Interpretive phenomenological analysis

**DOI:** 10.1101/2023.05.30.23290724

**Authors:** M. Ndu, G. Teachman, J. Martin, E. Nouvet

## Abstract

**Background:** The past three years have seen an increasing gap in health inequalities in Nigeria’s health systems, with many women having trouble accessing health care due to persistent social determinants of health. Studies indicate that the amplified impact of the pandemic is due to the lack of contextual focus on response plans. This study used an interpretive phenomenological analysis to analyze women’s experiences with healthcare as the pandemic progressed in Nigeria.

**Methods:** Semi-structured interviews were conducted between August to November 2022. It was supplemented with three focus group discussions with mothers. The mothers were purposively recruited for their experiences with health care during the pandemic. The analysis of the interviews followed the interpretive phenomenological analysis approach.

**Results:** Twenty-four women aged 15 to 49 years with children between 0 and 5 years participated in this study. These women reported mixed experiences during the pandemic, with many attributing positive health behaviours to the pandemic. Following analysis, four themes emerged: i) Influence of diversity of Healthcare Practices and Beliefs on health-seeking behaviour; ii) Unpacking Systemic Barriers to Seeking Timely and Appropriate Health Care Services; iii) Women’s fear of contracting COVID-19; iv) Socioeconomic Burden for Holistic Health Care Delivery.

**Conclusion:** Health planners must examine contextual factors that drive health usage, especially potentially changing gender dynamics ahead of the next pandemic. This paper examined women’s decision to seek or not seek care, the type of care they received, and where they went for care. Women felt that the pandemic affected their decision to seek or not seek care. However, while they learned new behaviours that are now integrated into their daily lives, they also indicate that some behaviours are habitual and have persisted through the pandemic.

## Background

Timely and appropriate services are critical to maintaining and sustaining the health of women and children. In the last decade, there has been a consistent focus on improving health services as a fundamental approach to minimizing mortality from preventable causes in Nigeria and the sub-Saharan region (1,2). Emphasis has been on pregnant and breastfeeding mothers, who constitute the highest proportion of global deaths from preventable causes, given the complicated cascade of events resulting in pre- and postpartum deaths (3). Nigeria, for instance, contributed 14% of the global maternal deaths in 2020, with over 58,000 women dying from pregnancy-related deaths (4,5).

In the last three years, the pandemic has also amplified existing inequalities in healthcare access and the need for more investment in health systems in sub-Saharan Africa. The impact of the pandemic on these healthcare systems is significant, with many countries facing shortages of essential medicines and equipment and health worker stress and burnout.

Like many other sub-Saharan African countries, Nigeria faces considerable impediments to women’s access to healthcare owing to factors such as poverty, limited education, and cultural norms prioritizing men over women (5–9). These barriers have been further compounded by the COVID-19 pandemic, which has resulted in disruptions in healthcare systems, economic downturns, increased household stress and burdens, and amplified disparities within and between nations (10–12). Such challenges have become more apparent in light of the pandemic’s one-size-fits-all approach, despite variations in the severity of its impact on individual countries (Cash & Patel, 2020). This realization underscores the crucial role of context-specific responses in any emergency intervention.

At the onset of the pandemic, Cash and Patel (12) argued that pandemic preparedness and response strategies must be tailored to local contexts, considering factors such as population density, access to healthcare, and social determinants of health. They also emphasize the importance of community engagement and participation in pandemic response efforts, as well as the need for global cooperation and solidarity to ensure equitable access to vaccines and other essential health resources. Therefore, it is crucial to redefine the equity agenda of global health beyond analyzing determinants of COVID-19 disease to also include scrutiny of the short and long-term effects of public health measures implemented in response.

This paper is a component of a larger study which aimed to analyze women’s experiences with healthcare in three states of Nigeria during the pandemic, as these experiences have implications for future pandemic preparedness, planning, and national health policy. By examining where women sought healthcare during the pandemic and their reasons for doing so, this paper sheds light on the implications of the pandemic for women’s health in Nigeria and globally. This paper can help identify and address healthcare barriers that women face in sub-Saharan African countries like Nigeria, such as poverty, lack of education, and cultural attitudes. Recognizing how such factors intersect with women’s health-seeking behaviour is important for promoting health equity, a fundamental principle of global health (13,14). Understanding how women accessed healthcare during the pandemic can inform future pandemic preparedness and response efforts. This knowledge can be used to address gaps in access and ensure that women’s health needs are addressed during emergencies.

## Method

### Study Setting

This research was conducted in targeted rural and semi-urban communities located in three Nigerian states, namely Sokoto (northwest), Ebonyi (southeast), and Ogun (southwest). The selection of these three states provides an opportunity for a comprehensive analysis of various social and cultural factors that may shape health-seeking practices. The inclusion of these particular states was purposeful and aligned with the research objectives, which strived to ensure the representation of diverse experiences of women from distinctive regions in Nigeria. We did not aim to generalize findings across all Nigerian women but rather to facilitate an understanding of potential social and cultural influences on healthcare-seeking behaviours among women navigating infectious diseases outbreaks in culturally distinct states. The study locations had a primary health care centre, with local pharmacies and private hospitals, except for Sokoto, which only had a primary health care centre. The services available to women through these sites ranged from routine services, i.e., malaria treatment and immunization, to delivery. The private hospitals also offered surgical services (i.e., Caesarean section).

### Study design

Interpretative Phenomenological Analysis (IPA) was employed as the research methodology in this study. IPA is a qualitative research approach that explores participants’ perceptions and subjective experiences in depth(15). As an interpretative approach, IPA recognizes that researchers’ interpretations play a vital role in the analysis process, resulting in what is referred to as a double hermeneutic (16). Thus, IPA aims to produce in-depth interpretative accounts of a small number of participants rather than a generalizable account for a larger sample, as it recognizes that each participant’s experience is unique and cannot be generalized to others (17). IPA’s interpretative and exploratory approach is particularly useful when examining complex and subjective experiences, such as women’s experiences with healthcare during the COVID-19 pandemic in Nigeria. By focusing on a small number of participants and producing rich and detailed accounts of their experiences, IPA can provide valuable insights into the lived experiences of these women and help identify areas where healthcare services can be improved to meet their needs better.

Towards answering the research question, “Where and why did women in Nigeria seek or delay health care for themselves or their children during the COVID-19 pandemic?,” we asked participants about their behaviour during the pandemic when their child was sick, focusing on the period spanning the lockdown and total and partial restrictions of interstate and intrastate travel. As such, we designed in-depth interviews and focus group discussion guides with open-ended questions to generate information on women’s experiences with healthcare-seeking at different temporal points -- before the pandemic, during the restriction of movement from March 2020 to July 2020 and after the restriction of movement. These guides invited women to recall details of their healthcare-seeking decisions in each of these periods, elaborate on factors they identified as influencing if, when, and where they sought care, and share details of their encounters with healthcare services if these were sought out. Before the study commenced, the lead researcher piloted the tool in one study location to refine and ensure the prompts were clear and understandable. All in-depth interviews and focus group discussions were conducted in the dominant local language of the data collection site, including Igbo, Hausa, and Yoruba. The tools were also translated using professional language translators from the Nigeria Translators Association. The full interview guides are available as supplementary materials.

### Participants

With the assistance of the local partner, Jhpiego Nigeria, women were recruited through the local women’s groups in the community. The research assistants who were natives of the study location were the local mobilizers of the women in the communities. Some women recruited for the focus group discussion indicated having had a health emergency during the pandemic. In contrast, others confirmed they had no health issues during the period under study. However, participants had to have a child aged 0-5. We sought to hear from women who gave reasons both for seeking and not seeking care during these periods. Given the significant barriers to accessing healthcare, we were interested in women’s perspectives. There was no requirement for women who participated in the focus group discussions to participate in the in-depth interviews. As such, recruitment to the in-depth interview followed a snowballing approach, where women who attended the focus group discussions informed their friends and neighbours about the study. These women were provided with the mobile numbers of the Research Assistants whom they contacted to indicate interest in participating in the interview. Following this contact, the Research Assistants and the lead researcher fixed a date and time to meet with the women at a convenient place.

Women were recruited as participants until 24 interviews were completed. In using IPA, Smith et al. (15) recommend that researchers use a small sample size given its idiographic commitment. Smith and colleagues, while noting that there is no correct answer to the question of sample size in IPA, recommend three participants for undergraduate and master’s level IPA research and 4-10 participants are recommended for a higher-level degree. Our decision to recruit a larger sample size was guided by our aim to generate rich and compelling data that represented multiple perspectives within a heterogeneous population. Given that data collection occurred across three sites, detailing some women’s personal experiences, we feel this number is small to make inferences on the broader Nigerian women based on the finding because of the diversity of women in the study context.

### Data collection

The lead researcher and the three research assistants conducted the focus group discussions with alternating roles as note taker and recorder. After each focus group, the lead researcher and the research assistants debriefed on observations of the non-verbal information from each session which the lead researcher recorded in her filed notes. Each focus group discussion lasted for approximately an hour and thirty minutes. The twenty-four interviews and three focus group discussions of 10-12 women each were conducted between August and November 2022. The first author was responsible for the interviews with the support of three Nigerian research assistants with prior qualitative research experience. Each interview lasted approximately 45 to 60 minutes. The team used eight open-ended questions as a guiding structure for the interviews.

### Data analysis

After the interview and focus group discussions, the research assistants, all indigenes of the data collection sites, transcribed the transcripts. The lead researcher working with a translator transcribed and reviewed the transcripts against the audio for accuracy. According to Smith et al. (15), researchers conducting Interpretative Phenomenological Analysis (IPA) are advised to fully immerse themselves in the interview transcript, reading each interview multiple times to comprehend the verbal expressions of the research participants. In line with this recommendation, the lead researcher initiated the data coding process by meticulously reading and re-reading each interview, carefully documenting any novel ideas and insights using coloured notes. The coding process was carried out manually, per the guidelines of Larkin and Thompson [18], emphasizing reflexivity to facilitate the researcher’s exploration of her presumptions, preconceptions, biases, and emotional responses. Through the repeated reading of each interview, the lead researcher identified codes and patterns of response, critically assessing her interpretations and returning to the recorded interview where necessary to ensure an ethical representation of the emotions and expressions of the participants. Subsequently, the lead researcher collated the themes and subthemes that emerged from the coding process, creating a comprehensive framework that captured women’s experiences. Throughout the analysis, the principal investigator engaged in ongoing discussions with EN, JM, and GT, ensuring that the themes and subthemes were traceable to the participants and that emerging patterns were systematically identified and recorded. This methodological approach was undertaken to strengthen the overall credibility of the study (18).

## Ethical approval

Ethical approval was obtained from the Western University Health Sciences Research Ethics Board (HSREB) reviewed and approved under the Project ID: 120417 and Review Reference: 2022-120417-70507, National Health Research Ethics Committee (HREC) Approval Number NHREC/01/01/2007-14/04/2022, at the Federal Ministry of Health, the Sokoto State Health Research Ethics Committee approval reference number SMH/1580/V.IV, Ebonyi State Health Research and Ethics Committee approval number EBSHREC/01/06/2022-02/06/2023, and Ogun State Health Research Ethics Committee approval reference number HPRS/381/464. No identifiers were used during the interviews and focus group discussions while transcribing to maintain the privacy of the women. Participation in the study was voluntary, and women knew they could decide at any point to opt out. The women provided both verbal and written informed consent during the interviews and focus group discussions.

## Results

The study sample comprised women aged 15 to 49 years with children aged between 0 and 5 years. A total of 24 women with diverse educational and socioeconomic backgrounds participated in the study. Specifically, 13 women had completed some secondary education, five had a college education, and six had obtained a university degree. Of the 13 women who had completed secondary education, eight were not employed. The remaining 18 women held various occupations, including teaching, catering, community health work, and civil service. The women were from different socioeconomic backgrounds, with some facing constraints to meet their basic daily needs. Most of the women were married, with only two exceptions: one a divorcee and the other a widow. Women lived at various distances from the closest hospitals and primary health care centers. While several women reported travelling for up to an hour to access the nearest referral hospital, many reported living within walking distance of the nearest primary healthcare center. Women in the study reported using various healthcare resources, including professional contacts and traditional remedies. Of the 24 women interviewed, 20 had some form of interaction with the health system during the pandemic. At the same time, the remaining four indicated they had no reason to visit any hospital.

**Table 1:** Participants characteristics.

|  |  |  |
| --- | --- | --- |
| Age |  |  |
|  | 15-30 | 14 |
|  | 31-45 | 9 |
|  | 36-49> | 1 |
| Education |  |  |
|  | Secondary school | 13 |
|  | University | 6 |
|  | College, i.e. HND | 5 |
| Employment |  |  |
|  | Paid employment | 6 |
|  | Self-employed | 10 |
|  | Unpaid employment | 8 |

In what follows, we present four interconnected key themes that emerged from women’s accounts of their health-seeking behaviours during the pandemic: i) Influence of diversity of Healthcare Practices and Beliefs on health-seeking behaviour ii) Unpacking Systemic Barriers to Seeking Timely and Appropriate Health Care Services iii) Women’s fear of contracting COVID-19 iv) Socioeconomic Burden for Holistic Health Care Delivery. Together these themes highlight an interplay between contextual conditions, norms, ideals, and women’s decisions of when, why, and from whom to seek healthcare during the pandemic. These findings reveal that where and why women in Nigeria sought or delayed health care for themselves or their children during the COVID-19 pandemic varied depending on the severity of the illness, social connections around their immediate environments, understanding of the pandemic, and personal beliefs about health in general. The four overarching themes from our analysis are presented using an interpretive approach.

### Theme 1: Influence of Diversity of Healthcare Practices and Beliefs on health-seeking behaviour

This theme sheds light on the diverse healthcare practices and beliefs within the community impacting healthcare-seeking behaviour, with some women relying on traditional or spiritual remedies while others seek professional medical services. Several norms and beliefs fall under this theme, each explained in detail below.

#### Factor 1: Negative perception of hospitals and apathy of health professionals

Several participants expressed a reluctance to seek medical attention until their symptoms became too severe to ignore. One described relying on sleep to relieve headaches and only seeking medical attention if their symptoms persisted and became unbearable. This hesitance to seek medical attention unless the situation becomes dire potentially creates a barrier to accessing healthcare services and highlights potential constraints on early detection and intervention.

Negative perceptions of hospitals also appeared to impact women’s decisions to seek medical care during the period.

> *“Like me, I don’t like hospitals. During the lockdown, I only had a headache. Usually, if I am having a headache (Ori fifo) I will sleep once I sleep, I am okay, if I now see that my body is not that okay, that it is getting too much, I will come to the hospital to get treatment.” [FGD, R10]*
>
> *“I don’t usually go to the hospital what did I want to go and do at the hospital, the COVID (Coro) we know that they just use it to take our money, and we get to the hospital, they will be doing different kinds of things. See, as children of God, we will not encounter sickness that we cannot handle.” [IDI 5, Businesswoman]*

#### Factor 2: Religiosity as a normative practice of health prevention and treatment

Other women mentioned that they do not give their children any medication, instead relying on the grace of God and prayer, which gives them the confidence to live normally, regardless of the presence of COVID-19. This suggests that some women in the community may rely on faith-based or spiritual remedies for their health, which may not always align with scientific or medical practices.

> *“I don’t give my children anything it is the grace of God and prayer water of Ayodele Babalola the water from Ikeji …Covid or no covid, we live our life normal.” [IDI 7, Businesswoman]*

#### Factor 3: Habitual practice of self-medicating

Several women indicated that self-medication is a common practice and described a cultural norm of self-medication in their village. They expressed that seeking medical attention from a hospital is not considered essential in their community and is only done when the illness is severe. While this approach may be reasonable, it can also be taken to the extreme when health-seeking behaviour is avoided or delayed due to prejudices against formal healthcare, even when the disease is severe or best served by formal care. This suggests a potential lack of awareness regarding the significance of preventive healthcare and early detection, which could negatively affect the community’s overall health. Most women acknowledged that self-medication was a practice they grew up with and did not see anything wrong with continuing the practice during the pandemic. This illustrates how their cultural upbringing and beliefs about health and healthcare practices have shaped their attitudes toward seeking medical attention. Since self-medication for minor ailments is rational and encouraged in all settings as it represents the most efficient and cost-effective management of most minor illnesses, it is the extreme and inflexible interpretation of self-reliance which may lead to harm and not the issue of self-medication that is itself to be questioned overall. However, the women’s depiction of self-medication as a favourable societal norm highlights the role of social pressure on amplifying the negative impact of internalized belief systems, which, when consistently repeated over generations, become an acceptable way of life:

> *“When growing up in the village, we normally do self-medication. No one is bothered about going to the hospital. We do not see it as something important. We only go to the hospital when it is severe.” [FGD, R6]*
>
> *“Truly if it is for a minor illness like a headache, I do not go to the hospital, but aside from that I do go to the hospital when my child is sick. But, then every time the child is ill we do not take them to the hospital because of fear of corona.”[IDI 23, No Paid Employment]*

#### Factor 4: Preference for local chemists

During the initial stage of the pandemic, restrictions on intra and inter-state movements compelled many women to rely on local chemists, which has been a common practice. Some women preferred consulting with a local chemist when seeking healthcare advice. The proximity of the chemist to their community and their success in receiving what they perceive as effective treatment from chemists in the past are significant factors in this preference. The same women who noted a preference first to seek care from the local chemist also noted that they eventually took their child to the health center for professional medical services. This extract highlights the importance of accessibility and affordability of healthcare services, which can impact women’s health-seeking behaviours.

> *“I first took him to the chemist to complain about what was wrong with him, so the chemist told me to take him to the hospital, I took him to the health center. But I still prefer the chemist a little. Chemist is close to us; once we get there and we complain about what is doing us when they write drugs for us, and we use, and we see changes, that is okay.” [IDI 8, No paid Employment]*
>
> *“The decision to go to the hospital depends on the severity of the illness. If the sickness lasts for three to four days and becomes complicated, we will go to the health center as quickly as possible. However, if it’s only a two-day sickness, we usually go to the chemist in our area for injections.” [IDI 15, Businesswoman]*

#### Factor 5: Norm of combining medical and traditional medicine

Several women in the study reported using herbal or a combination of traditional remedies and prescribed medication for their health concerns. These women often turned to traditional remedies before seeking professional healthcare services. They would go to the hospital if they did not see improvement after three to four days. This reliance on traditional remedies suggests a preference for seeking medical care from non-professional sources within the community. These findings emphasize the need for healthcare providers to address cultural beliefs and perceptions surrounding traditional remedies to improve healthcare-seeking behaviours and outcomes within the community.

> *“I used drugs and herbs both of them are working together for me. Usually, after three to four days if we do not see any improvement, we will quickly go to the health center but for now, we will use the nurse in our area” [FGD, R15]*
>
> *“Some pregnant women believe in those [traditional healers] that give them drugs in the house like those trado-medical they prefer to go there because they believe that herbs and root are created by God despite the health problems they face after taking some traditional medicines” [FGD, R20]*

The above factors highlight the varied attitudes of the women interviewed toward healthcare-seeking behaviour and the role certain norms and internalized beliefs play in their healthcare-seeking behaviour. During the pandemic, many women reported staying home and using home remedies or chemists to treat themselves or loved ones. Others reported resorting to traditional practices as an alternative to hospital visits. In both cases, however, the women who did describe staying home or seeking traditional practices also reported visiting hospitals when their illness or that of a loved one became unmanageable at home.

One woman employs the phrase “no one is bothered” to rationalize her decision to engage in self-medication, thus demonstrating cognizance of the existence of appropriate and acceptable healthcare approaches that warrant exploration. However, due to the social norm of this practice, she perceives it to be socially acceptable and continues to engage in such behaviour. For instance, a mother recounted her experience of providing home-based treatment to her child under the presumption of a malaria infection. However, as the condition deteriorated, she realized a need to seek professional help. At that point, she encountered the challenge of seeking medical attention during a restricted movement period. Another woman describes treating headaches with sleep and only seeing a health professional when it becomes severe.

The women use words like “just”, “minor” and “severe” to indicate their perception of urgency for care. For instance, one woman uses the word “just” to describe headaches which suggests that it is not considered an illness that requires health practitioners’ attention. The women display the same disposition towards ailments, particularly those resembling symptoms of COVID, which evinces a reluctance to seek health care in such situations. To gauge the severity of an ailment, the women use the duration or length of an ailment as a metric. They downplay the severity of their ailments using “minor” to describe their unwellness and lessen its importance while using “severe” to describe that of their children. The women also seem to indicate a lack of awareness that the manifestation of symptoms like headaches and fever could suggest an alternate underlying ailment requiring an accurate diagnosis before administering treatment. This highlights the potential for women to adopt unsafe practices when restrictions prevent them from accessing professional healthcare.

### Theme 2: Unpacking Systemic Barriers to Seeking Timely and Appropriate Health Care Services

This theme explores the systemic factors that influenced women’s decision-making regarding healthcare-seeking behaviours during the COVID-19 pandemic. More specifically, it presents the various barriers within the health ecosystem women reported encountering, ultimately impacting their willingness to seek care. The following are the factors contributing to this theme:

#### Factor 1: Potential fear of COVID-19 (mis)diagnosis

This factor highlights how fear of COVID-19 may prevent women from seeking necessary medical attention, even for non-COVID-related illnesses. It also includes the emotional and psychological impacts of the COVID-19 pandemic beyond just the physical health consequences, underscoring the importance of continuously educating the public about infectious disease symptoms and addressing misinformation through health literacy campaigns.

Some pregnant women felt that if they sought out healthcare from a hospital, they might be subjected to stigma and misunderstanding due to their pregnancy symptoms, which can be mistaken for signs of COVID-19. They reported feeling weak and experiencing sneezing, which was sometimes misinterpreted by others as signs of the COVID-19 virus. These perceptions could suggest an internalized stigma from past experiences dealing with health workers and society during epidemics or other infectious disease outbreaks, including Ebola virus disease. This suggests the potential for stigma and discrimination towards pregnant women due to misunderstandings of their condition.

> *“During pregnancy, we experience symptoms that can make us feel weak, and sometimes people mistake that weakness for a sign of COVID-19. Similarly, if we sneeze, some people assume we have the virus.” [IDI 17, Businesswoman]*

Linked to the above expressed concern about being subjected to stigma and discrimination, potentially, some women reported that the first thought that came to mind when feeling ill was whether they have the COVID-19 virus. This fear may lead to social isolation and loneliness, as women may avoid going out and interacting with others. This highlights the emotional impacts of the pandemic on women beyond just physical health consequences.

> *“When you start feeling sick, the first thing that comes to mind is whether you have COVID-19. You don’t want to go out and have people say that you have it.” [FGD, R20]*

Other women described the impact of the pandemic on medical professionals’ willingness to provide care. Some reported that at the onset of the pandemic, doctors were hesitant to touch patients, especially those with symptoms related to breathing or chest pain, cough, or sore throat, due to fear of the COVID-19 virus. This could have negative consequences for those needing medical attention if they feel the health provider is reluctant to engage in a familiar manner.

However, participants also noted a decrease in fear and stigma associated with COVID-19 as the pandemic progressed, and people understood the virus better, suggesting potential positive changes over time.

> *“When the COVID-19 pandemic started, doctors were hesitant to touch patients, especially if they had symptoms related to breathing or chest pain, or if they had a cough or sore throat. They were afraid it could be COVID-19. But thankfully, we haven’t heard much about the virus lately.” [IDI 4, Businesswoman]*

In the below interview extract, the woman expresses fear of taking their child to the hospital during the COVID-19 pandemic due to concerns that their child might be diagnosed with COVID-19. Instead of taking the child to the hospital, she opted to visit a chemist.

> *“During the COVID-19 pandemic, my child had a cold, but I was afraid to take him to the hospital, fearing that they would say it was “coro,” so I took him to a chemist. After two days, here was no improvement, so I took him to the hospital, and they said he has catarrh [excessive buildup of mucus in the nose or throat] and typhoid”* (FGD, R4)

Other women mentioned that they do not go to the hospital for minor illnesses like headaches but will seek medical attention when their child is sick. However, they also mention that fear of COVID-19 prevents them from taking their child to the hospital whenever they are ill.

> *“Truly if it is for a minor illness like a headache, I do not go to the hospital, but aside from that I do go to the hospital when my child is sick. But, then every time the child is ill we do not take them to the hospital because of fear of corona.”[IDI 23, No Paid Employment]*
>
> *“During COVID-19, anytime I am sick, I used to be afraid of going to the hospital so that they would not say I have coronavirus. So, I take Panadol.” [FGD, R6]*
>
> *“One of the major challenges was visiting a doctor whenever I have a cold or Catarrh [excessive buildup of mucus in the nose or throat] because of fear of being diagnosed with COVID-19” [IDI 20, Businesswoman]*

#### Factor 2: Difficulty using public transportation during pandemics

This factor highlights the challenges women face in accessing healthcare services during the COVID-19 pandemic, particularly in areas with limited transportation options. The extracts also demonstrate the importance of community support and safety measures in ensuring access to healthcare services for vulnerable groups such as pregnant women and children.

One mother described her experience with her child’s illness during the pandemic. Due to movement restrictions, they could not leave the house, and the child’s condition continued to worsen despite initial treatment with anti-malaria medication. As there were no available vehicles in their area, they had to walk to the hospital, which was a strenuous journey. However, they were fortunate to encounter someone who offered them a ride. The experience highlights women’s challenges in accessing healthcare services during the pandemic, especially in areas with limited transportation options. It also emphasizes the importance of community support in overcoming such challenges.

> *“During the COVID-19 pandemic, one of my children came to spend the vacation with me and fell ill. Due to the restrictions on movement, we were unable to leave the house, so I had to call [on mobile] on health workers to assist. Initially, I treated him with anti-malaria medication, but when he continued vomiting, I realized we needed to take him to the hospital. Unfortunately, there were no available vehicles in our area, so we decided to walk to the hospital. It was a strenuous journey, but we were fortunate to encounter someone who was allowed to be out during the lockdown and who kindly offered us a ride. We eventually arrived at the health center where the child received good care.” [IDI 7, Businesswoman]*

A pregnant woman discussed her experience during the pandemic taking a taxi to her antenatal appointments as she did not have a car. However, the experience was challenging due to the close proximity of passengers in the taxi. She had to pay for two seats to avoid touching other people. This highlights the fear and anxiety pregnant women face during the pandemic regarding exposure to the virus. It suggests the need for transportation alternatives and safety measures to address pregnant women’s challenges in accessing healthcare services during the pandemic.

> *“Ehmm, for me, the challenge was that when I went to ante-natal because we did not have a car, I had to take a taxi, and you know how those drivers usually pack us like sardines [tight]. I always pay for two seats, so I do not touch my body with people.” [IDI 7, Student]*

#### Factor 3: Trust and Familiarity with Health Providers influence where women will seek care

This factor highlights the importance of having a strong relationship with a healthcare provider, especially in communities where access to healthcare may be limited. One woman mentions that she went to a private doctor whom she knows and who trained her as an auxiliary nurse when she had her baby. The doctor gave her drugs and some money as they did not have them. This suggests that the nurse has a strong relationship with the doctor and trusts him to provide her with medical assistance. She also mentions that she always calls him when they are sick, indicating that he is her go-to person for medical assistance. As such, she relied on him during the pandemic when they were sick.

> *“At the time, I had my baby. I went to private because I know the doctor. He is the one that trained me as an auxiliary nurse. I called him [on mobile], and he said to come. He gave us drugs and some money because we did not have it. I always call him when we are sick.”[IDI 24, Trained Auxiliary Nurse]*

Similarly, another woman mentions that she lives on the road opposite the community center, and her friend, whom she refers to as “Aunty Nurse,” is always available to provide medical assistance when they are sick. Even when she does not have money, the nurse will write drugs for her. This highlights the close relationship between the two women and their trust in each other.

> *“I live on the road, opposite the community centre. Aunty Nurse is my friend and answer when I call [mobile]. That time if I or anyone in my house is sick, we just go to her. Even if I don’t have money, she will write drugs for us” [IDI 9, Business Woman]*

#### Factor 4: Mistrust of the healthcare system

This factor underscores the significance of examining the larger societal and cultural context in which women experienced the COVID-19 pandemic and access healthcare. The interview extract features a woman sharing her perspective on health during the COVID-19 outbreak. Her statement suggests a perception of the pandemic and the role of medical professionals and politicians shaped by suspicion and mistrust. Specifically, she believes that medical professionals and politicians are collaborating to create problems for people. This viewpoint points to a broader societal issue of mistrust in authority figures and the system, which may impact healthcare-seeking behaviours during the pandemic.

Additionally, the woman’s belief that good health prevails when things are running smoothly highlights the significance of stability and predictability in maintaining health. This perspective contrasts with the uncertainty and disruption created by the pandemic, which can increase stress and anxiety that negatively impact health.

> *“My view regarding health during the Covid (Coro) outbreak is that medical professionals are colluding with politicians who seek to create problems for us. Therefore, I believe that when things are running smoothly, good health will prevail among us.” [IDI 4, Businesswoman]]*

The factors above underscore the significance of comprehending patients’ experiences and perspectives when implementing effective healthcare interventions, particularly in relation to pandemics and outbreaks. The women’s fear of COVID-19 and discomfort with health workers’ PPE initially influenced their decision to seek alternative treatment. The women articulated their challenges in adhering to COVID-19 prevention measures when accessing healthcare. However, their persistence in seeking appropriate medical care ultimately led to an accurate diagnosis and treatment. The factors also suggest that misunderstandings and internalized stigma may contribute to poor emotional health for pregnant women. The women included in this study used words like “weak” and “weakness” to describe their perception of the physical state of pregnancy. It indicates an internalized ethical or moral judgment of themselves in pregnancy and how it presents to others. It also shows that women value the opinion of others when ill and the emotional or psychological stress such needs can place on women, particularly during pregnancy, as they do not want to be condemned for being in public while sick. For instance, one woman describes being distressed about going out in public when coughing, as onlookers may assume she has COVID.

For women who struggle with such internalized stigma, this indicates they will go into stigma isolation to avoid unpleasant experiences. The factors indicate the importance of addressing stigma and misinformation related to infectious diseases and the need to support women who may experience stigma isolation due to pandemics. The trust, distrust, and familiarity women share with their health providers and the health system broadly play critical roles in ensuring their willingness to seek care. Additionally, it highlights the need for healthcare providers to establish trust with their patients to ensure that they receive the best care possible. It also stresses the importance of healthcare professionals establishing trust and building rapport with patients to enable effective communication and treatment.

### Theme 3: Discomfort with COVID-19 protocols

This theme highlights women’s perception of COVID-19 protocols and the impact on their healthcare-seeking during the pandemic. It presents some challenges navigating the health system preventive measures, including complying with the instructions given by healthcare providers, following the procedures put in place by hospitals, and discomfort with certain aspects of healthcare services. It underscores the importance of public health messaging and education to dispel fears and encourage women to seek medical attention while also following COVID-19 protocols to ensure their safety.

Some women expressed that receiving instructions from nurses to comply with COVID-19 prevention measures, such as wearing masks and washing hands, was distressing and bothersome. However, they also recognized that complying with these measures was necessary. This indicates that the women recognize the significance of adhering to these measures to prevent the spread of the virus but may be experiencing some emotional or psychological discomfort while doing so.

> *“The nurses are always bothering us with instructions to wear nose masks, wash hands and do this and that. It can be stressful, but we have no choice but to comply.” [IDI 18, No Paid employment]*
>
> *“Sometimes, going to the hospital can be difficult due to the procedures they have in place. They ask us to keep a certain distance from others, wash our hands, and wear a face mask before entering. It’s not easy to comply with all of these rules.” [IDI 19, No Paid Employment]*

Other women reported feeling uneasy with how nurses during the pandemic must wear gloves before attending to them to either check blood pressure or administer injections. This suggests that the nurses’ conduct may be off-putting, indicating a potential lack of trust or comfort with the healthcare provider. Such negative experiences may influence some women’s willingness to comply with other healthcare instructions, including those related to COVID-19 prevention. When queried if this experience was unique to COVID-19, the women mentioned that health providers stopped using gloves while attending to them since the relaxation of restrictions.

> *“When they check our ifunpa giga [blood pressure], they wear gloves, but when it comes to injections, we don’t like the way they do it. They usually act in a certain way that puts us off.” [FGD, R5]*

They acknowledged that these measures were implemented for safety reasons but also highlighted the difficulties of following them, such as maintaining physical distancing, wearing a face mask, and using PPE. This signifies that the COVID-19 pandemic has influenced healthcare experiences beyond the direct impact of the virus, and healthcare providers may need to consider the additional burdens imposed on patients in implementing prevention measures.

### Theme 4: Gendered notions of responsibility and its effect on health-seeking

This theme explores women’s decision-making power to secure funds needed for timely care and their role in the family. It also suggests that health care in Nigeria incurs financial pressures, and women still need to secure funds for health in the absence of social safety nets. The women all mentioned their husbands and their primary role in financial decision-making.

> *I have to tell our husband because it is mandatory, I do so since he is the head of the house. We asked him if he had money to give me he does, but if not, he would suggest how to get it. [IDI 24, Businesswoman]*
>
> *“My husband is a pilot (tricycle driver), he is trying. He will give me the money because my business is small and I was not selling anything then. My fish business is very small. It is our children.” [IDI 10, Businesswoman]*

Other women reinforce the idea that husbands are often viewed as the primary financial providers in the household, and women may rely on them for financial support. Several women described finding the funds necessary to seek health care for a child when their spouse is absent.

> *During that time, my child was sick, and the father travelled then, so taking care of myself was difficult. I borrowed money from my neighbour and then paid it back when the father returned. [IDI 20, No paid employment]*
>
> *“Why would I not tell my husband? He is my oga na! Ah, if my child is sick, I will tell him the child is sick, even if he is travelling, I will call him and tell him. He will ask me to borrow the money, and he will pay when he comes back. I will go to my brothers, they will always give me.” [IDI 13, Businesswoman]*

Some women expressed that they would only borrow from close family members. They felt that often asking for help from people outside of their immediate family would reduce their social image. More specifically, women felt that asking outsiders for help, including friends, could result in petty gossip, which in small communities tends to be stressful.

> *“That time, people did not have. I am the sole breadwinner for my family. My husband did not have a job because, during the covid, nobody was digging boreholes. I always ask my siblings, but if they do not have, I will manage. [IDI 16, Businesswoman]*
>
> *“If my husband does not have to give, I will ask my mother and my sisters, if they do not have, I will not ask my friends or neighbours. You know how people talk. Tomorrow they will say, so and so is suffering and is even borrowing money from everyone.”[IDI 12, Auxiliary Nurse]*

The above interview extracts centred on the financial practices of women who engage in various forms of work, whether paid or unpaid. The women in question commonly conveyed a relinquishment of financial control to their husbands, who they identified as the heads of their households. Some women with business ventures or paid employment seemed to downplay their financial independence, suggesting that women in such circumstances may intentionally, in accordance with cultural norms, stress their reliance on their husbands for financial support and that men are traditionally seen as the primary breadwinners in the household. It is noteworthy that societal expectations can further exacerbate the situation, with some women feeling pressured not to spend their own money, thereby restricting their financial autonomy.

Furthermore, some women expressed a sense of unease at seeking financial assistance beyond their immediate family members. In contrast, others were open to exploring all potential avenues for aid in support of their families. For instance, one woman remarked that if her husband could not provide for their needs, she would instead turn to her mother, siblings or neighbours, and friends. Such perspectives raise critical questions about the accessibility of healthcare services, both during and outside public health emergencies, since healthcare in Nigeria requires out-of-pocket payment and imposes an additional burden on women. Not only does healthcare pose a financial challenge to many families, but women responsible for organizing any needed healthcare for their children must consider how their decision-making processes and healthcare financing can uphold or tarnish their reputation, marriage, and household reputation based on gendered and social norms.

## Discussion

This study aimed to understand women’s behaviour while seeking care during the pandemic and where and why they chose to seek or not seek care. These findings contribute to a growing body of literature on women’s experiences during the pandemic’s early days and, in general, women’s experiences accessing health care during infectious disease events (12,19–35) To the best of our knowledge, this is the first interpretative phenomenological analysis of these experiences in Nigeria.

At the height of the current pandemic, early predictions estimated a negative impact of the pandemic on women’s health-seeking behaviour in Nigeria and globally, with eroding health gains for mothers and children in the last three decades (33,36). A particular concern noted in the literature was women’s ability to access quality care promptly and the access to financial resources to facilitate access (20,34,37). According to the WHO (30), the disproportionate impact of the pandemic on women increased gender inequities, threatening women’s empowerment and well-being in many countries, including Nigeria. While pandemic restrictions have lifted in most contexts, service availability and use of routine services in many countries remain disrupted (38).

In this study, our results demonstrate that during the pandemic, women experienced barriers to seeking or accessing healthcare for themselves and their children due to fear of contracting COVID-19 at health facilities or in transit, financial constraints, and lack of transportation. With the lockdown and various restrictions on movement, women had to look for alternative means to access care. Our findings indicate that women leveraged technology to speak to health workers who were relatives or friends. Many of them expressed that the geographic distance from health workers did not affect their ability to seek healthcare. Although, these women did express that self-medicating was their first choice before reaching out to health workers. The use of technology, such as mobile phones, by women to communicate with friends or relatives of health workers highlights the importance of technological innovations in improving healthcare access and delivery. This finding shed light on the complex interplay between individual healthcare-seeking behaviour, technological innovations, and external factors such as pandemic-related restrictions, highlighting the need for context-specific and multi-faceted approaches to improving healthcare access and delivery for women.

Some women reported attempting to seek hospital treatment only when their illnesses became unmanageable at home. This describes a pattern of self-medicating behaviours that, women participants asserted, existed before the COVID-19 pandemic. Nonetheless, our results indicate that pandemic-related restrictions on mobility and lockdowns likely exacerbated these behaviours. In some cases, women felt compelled to self-medicate or call friends and relatives in healthcare for mobile consultations due to limited options for hospital treatment resulting from movement restrictions and lockdowns, with a few encountering difficulties securing transportation to hospitals, often resorting to travelling significant distances on foot.

Additionally, religion and spiritism were reflected as alternative preventive and treatment measures for women who believed the pandemic was an illness only treatable by prayer. In Nigeria, as in numerous other African nations, religion and spirituality are deeply embedded in people’s daily lives, constituting a significant factor in the perception and management of health-related issues. This cultural orientation has played a vital role in guiding the response and coping mechanisms of the people to the challenges presented by the COVID-19 pandemic. This belief may be linked to early religious predictions on the outcome of the pandemic touted by religious leaders in Nigeria who refused to adhere to the restrictions and subsequently (39–42). This finding contradicts a study in southwest Nigeria which reported that while spiritism influenced health-seeking behaviour, participants did not perceive the pandemic to be an act of God (42). This attitude towards the pandemic was not unique to women in Nigeria. Globally, there was evidence of a religious stance against COVID as an illness not meant for “true” believers of God (43).

From women’s perspective, culturally assigned social roles for men and women gave men the role of head of household and, as such, defined them as the providers. This finding revealed that women tended to defer to their husbands for money and permission, even when they had their own money. Women’s perception of their own role in the family as one of support which ultimately does not include financing family expenses. Many women expressed concerns about letting their spouses know they have money, as it may reduce their spouse’s financial commitment to the house. Their perception illustrates a deep-rooted cultural norm around gender that explicitly positions men and women as providers and caregivers, respectively. With the advent of globalization, more women are becoming financially empowered in Nigeria (44). Yet, from the findings, it is evident that many women still see their role as support figures in the family.

The social expectation of women to continue to maintain their traditional roles, regardless of the financial contributions they make to the family, could be driving this attitude among women.

Social and cultural change can occur at different rates, with some changes happening quickly while others take much longer to take hold (45). While attitude to some social and cultural norms has improved over the years, in countries such as Nigeria, some changes have been slow (Varnum& Grossman, 2017). Several studies have found that the cultural role of men as breadwinners and women as caregivers have affected the division of labour and resources within households despite the influence of globalization on modern women. (46–48). Evidence indicates that social and cultural norms are slow to change. Women who do not have paid employment, while perceiving their role as supporters, would source for finance from friends, family and sometimes neighbours, while some other women felt that taking financial issues out of the immediate family circle could result in reduced social status in the community.

In Nigeria, the notion that the government is manipulating the public for political gain is deeply entrenched within the country’s social fabric. As governments attempted to persuade people about the severity of the COVID-19 pandemic, women may have perceived a conflict between the messaging and the general attitude of those they regarded as authority figures. The history of distrust between citizens and politicians in Nigeria (49–51) could be a direct reflection of the general distrust of the health system among citizens. This distrust of politicians, who frequently serve as the face of pandemic responses, may result in a lack of confidence in the health information dispensed by the government and a reluctance to seek necessary healthcare services. Women’s health, including access to reproductive and maternal healthcare services and increased susceptibility to infectious diseases, could be adversely impacted as a result.

Additionally, given the history of widespread mainstream media manipulation in Nigeria, people continue to turn to social media for information mainly due to their distrust of mainstream media, which they would perceive to be government sympathizers (52,53). The volume of misinformation penetrating communities from such social platforms has been shown to have an adverse effect on people’s reactions to the pandemic. According to several studies, one of the identified issues is the velocity of misinformation and disinformation circulated at the onset of the pandemic (19,54–56). Much of this misleading information was reiterated over digital media, with many religious leaders misleading congregations on its actual intensity. The framing of the COVID-19 messaging may have also affected how women received it. For instance, studies found that early messaging on the origin and source of the pandemic affected the perception of severity globally (19,55).

Women perceived health workers’ struggle in providing services while keeping themselves safe. The burden on the health system at the onset of the pandemic has continued to impact the Nigerian health system. The Nigerian health system is estimated to have 1.95 health workers (physicians, nurses, midwives) per 10000 people, with the highest concentration in urban centers. This is significantly lower than the WHO threshold of 4.45 health workers per 1000 people required to deliver essential health services and highlights the extraordinary burden on health workers in Nigeria to provide health services with insufficient human resources (57,58).

## Future Research

Future research could explore the impact of globalization on gendered and social roles in Nigeria. The findings from this study have implications for health planning and programming and feminist advocates. Studies should also examine women’s experiences with a disability during the pandemic. Evidence shows that women with disability face multiple barriers to accessing safe, respectable healthcare(59–62). It could also examine the role of massaging on women’s health literacy and health-seeking behaviour. This topic is essential as it highlights the importance of transparency in promoting health behaviours and addressing health disparities.

Additionally, women’s ability to leverage technology during this period indicates the potential of telemedicine in increasing access to healthcare. Given this, there would be merit in a study exploring the role of digital technology in increasing access to healthcare for women during pandemics. Finally, it is essential to examine health workers’ experience during the pandemic, given the increasing burnout and stress. With the global shortage of health workers estimated to continue over the long term, it is essential to understand the needs of health workers during future pandemics and other shocks to the health system.

## Strengths and Limitations

This study contributes to existing reports on women’s health experiences and perceptions of their health-seeking behaviour during infectious disease outbreaks and pandemics. Participant recruitment was done through the lead researcher’s professional network via a partner organization, Jhpeigo Nigeria, which also recommended the research assistants used in the study. All the research assistants employed in the study already have established relationships with the communities where the fieldwork for this study happened; as such, these may have influenced the dynamics and findings of the result as participants may give responses that they felt would be acceptable to them. Additionally, the lead researcher as an outsider may have affected the study results as participants may have withheld vital information that could provide robust data for analysis. However, the lead researcher switched roles with the research assistants once this was noticed to avoid any potential data contamination.

To further mitigate the risk, the lead researcher kept a field journal which included daily reflective diary entries concerning field activities. The journal supported the lead researcher in reflecting on the information shared by participants critically and actively. It was also helpful as a reflective tool for the lead researcher to examine her pre-existing biases, pre-conceived beliefs, and assumptions and acknowledge her positionality better to understand the phenomenon of women’s health-seeking behaviour and provide a thoughtful and nuanced interpretation. While the lead researcher piloted the tool, because of time and associated cost, the study tools were only piloted in one location. Given the heterogeneous nature of Nigerian society, it would have been advantageous to pilot the study tool in the three geopolitical zones of the study location. This would have ensured the clarity and appropriateness of the tool for study participants in the various locations. However, given the time and financial constraints, this was not possible.

## Conclusion and recommendations

This paper explored where women accessed health care during the pandemic and the mediators of their health-seeking behaviour. While this paper focused on where women accessed healthcare during the pandemic, it also explored the underlying reasons for their health-seeking behaviour and the specific types of care they received. By doing so, the paper provides a comprehensive understanding of the factors influencing women’s access to healthcare during this challenging time. The women felt that the pandemic influenced their decision to seek or not to seek care. While the pandemic may have influenced their decisions, the findings reinforce that certain habitual behaviours persist despite women knowing the implications of such behaviours. They recognized the challenges resulting from the pandemic, humanizing health workers’ experiences as needing care and support. The results support the need to improve health messaging and be transparent about the causalities and effects of diseases, recognizing that providing information increases acceptance of interventions.

Recognizing the enduring influence of religion and advocating with religious leaders to provide messaging that acknowledges and supports the separation of spirituality and access to health care. In policies and programming, it is crucial that physical healing is acknowledged as a biological or physiological process separate from spiritual healing. Overall, the COVID-19 pandemic has highlighted the importance of addressing the underlying social and economic factors contributing to women’s health-seeking behaviour in Nigeria and other resource-limited countries in sub-Saharan Africa. This includes investing in health systems and improving access to healthcare, especially for marginalized groups such as women and girls. We recommend that health planners consider the potential impacts of changing gender dynamics in this context since these considerably impact health planning.

## Data Availability

Following the Tri-Council Policy Statement: Ethical Conduct for Research Involving Humans policy, due to confidentiality agreements between participants and the research team, data cannot be made publicly available.

## Acknowledgements

The authors acknowledge the support of Jhpiego Nigeria, the Nigeria Federal Ministry of Health Ethics Committee, and the States Ethics Committees in facilitating the data collection and access to the communities.

